# SARS-CoV-2 in human sewage in Santa Catalina, Brazil, November 2019

**DOI:** 10.1101/2020.06.26.20140731

**Authors:** Gislaine Fongaro, Patrícia Hermes Stoco, Dóris Sobral Marques Souza, Edmundo Carlos Grisard, Maria Elisa Magri, Paula Rogovski, Marcos André Schörner, Fernando Hartmann Barazzetti, Ana Paula Christoff, Luiz Felipe Valter de Oliveira, Maria Luiza Bazzo, Glauber Wagner, Marta Hernández, David Rodriguez-Lázaro

## Abstract

We analysed human sewage located in Florianópolis (Santa Catalina, Brazil) from late October until the Brazil lockdown on early March. We detected SARS-CoV-2 in two samples collected independently on 27^th^ November 2019 (5.49±0.02 log genome copies/L). Subsequent samplings were positive until 4^th^ March 2020 (coinciding with the first COVID-19 case reported in Santa Catalina), with a SARS-CoV-2 RNA increase of one log (6.68±0.02 log genome copies/L). Our results show that SARS-CoV-2 has been circulating in Brazil since late November 2019, much earlier than the first reported case in the Americas (21^st^ January 2020, USA).

---

The first cases of atypical pneumonia related to severe acute respiratory syndrome coronavirus 2 (SARS-CoV-2) were described in Wuhan City, Hubei Province (China) on December, 2019 [1]. However, some indirect evidences could suggest that the virus had already been circulating a few months previously [2]. After the first reports, the number of cases increased exponentially around the world, and the WHO declared the status of pandemic on 11^th^ March 2020. As of 24^th^ June, 2020, SARS-CoV-2 has caused more than 9,1 M COVID-19 cases worldwide resulting in more than 473,000 deaths in 216 countries. The first diagnosed COVID-19 case in the Americas was reported on 21^st^ January 2020 in the USA (on 25^th^ February in Brazil) [1]. Since then, almost half of the diagnosed cases (49.4%) and deaths (47.8%) worldwide have been reported in that continent [1].

SARS-CoV-2 is a respiratory virus and the transmission route is mainly through respiratory droplets or contact with contaminated surfaces [3]. However, gastrointestinal and liver manifestations with high and prolonged faecal shedding of up to 10^8^ SARS-CoV-2 genome copies per g of stool have reported [4,5]. Consequently, the faecal-oral transmission of SARS-CoV-2 cannot be ruled out, although its role in COVID-19 epidemiology has not been yet determined [4].

The analysis of human and animal sewage has been successfully applied in monitoring the presence of biological risks [6], including enteric viruses [7]. A similar monitoring strategy has been applied for the detection of SARS-CoV-2 [8-15], demonstrating its value as a non-invasive early-warning tool for monitoring the status and trend of COVID-19 infection [12].

## Sampling urban sewage in Florianopolis, Santa Catalina, Brazil

We collected urban sewage samples in Florianopolis, Santa Catalina, Brazil (Figure 1) in 6 independent sampling events since 30^th^ October, 2019 until 4^th^ March, 2020. Raw sewage samples were collected from a separate collection sewage system in central Florianopolis, serving a population of approximately 5,000 inhabitants. The sampling was conducted using a submersible pump inside a well, used for inspection and cleaning of the network. The collection system receives wastewater exclusively, with a linear infiltration rate varying from 0.05 – 1 L / s Km, according to the Brazilian regulation for sanitation projects (ABNT – Associação Brasileira de Normas Técnicas n.9646/1986). In all the sampling events, 200 ml of urban sewage was collected and immediately transferred to the Applied Virology and Environmental Engineering Laboratory of the Federal University of Santa Catarina (Brazil), and stored at -80°C until its use. The concentration of the viral particles was performed using 25 mL of each sample as previously described [16]. Murine Norovirus (MNV-1) was artificially inoculated as sample process control virus to estimate the efficiency of extraction and the final viral load [17]. Viral RNA was extracted from concentrates using the QIAamp® Viral RNA Mini kit (QIAGEN, CA, USA) according to the manufacturer’s instructions, and eluted in 200 µL of RNAse free water. Viral RNA was detected by real-time RT-PCR (RT-qPCR) on a 7500 Real-Time PCR instrument (Applied Biosystems, USA). Sample process control virus, MNV-1, was detected as previously published [18]. SARS-CoV-2 was detected by OneStep qPCR Quantinova kit (QIAGEN, Germany) using oligonucleotides and probe previously published targeting the N1 [19], S and RdRp [20] regions of the SARS-CoV-2 genome. Oligonucleotides and a commercial SARS-CoV-2 RNA (2019-nCoV_N_Positive Control, 2 × 10^5^ genome copies/µL) as positive quantitative control were purchased from IDT (Integrated DNA Technologies, Belgium). Samples were considered as positive when the Cq values were ≤ 38.00. All RT-qPCRs were performed in duplicate in two independent experiments. All the positive samples were confirmed using the Seegene Allplex(tm) 2019-nCoV commercial kit in an independent laboratory (Laboratório de Biologia Molecular, Microbiologia e Sorologia, UFSC University Hospital, Florianopolis, Brazil).

**Figure 1:**
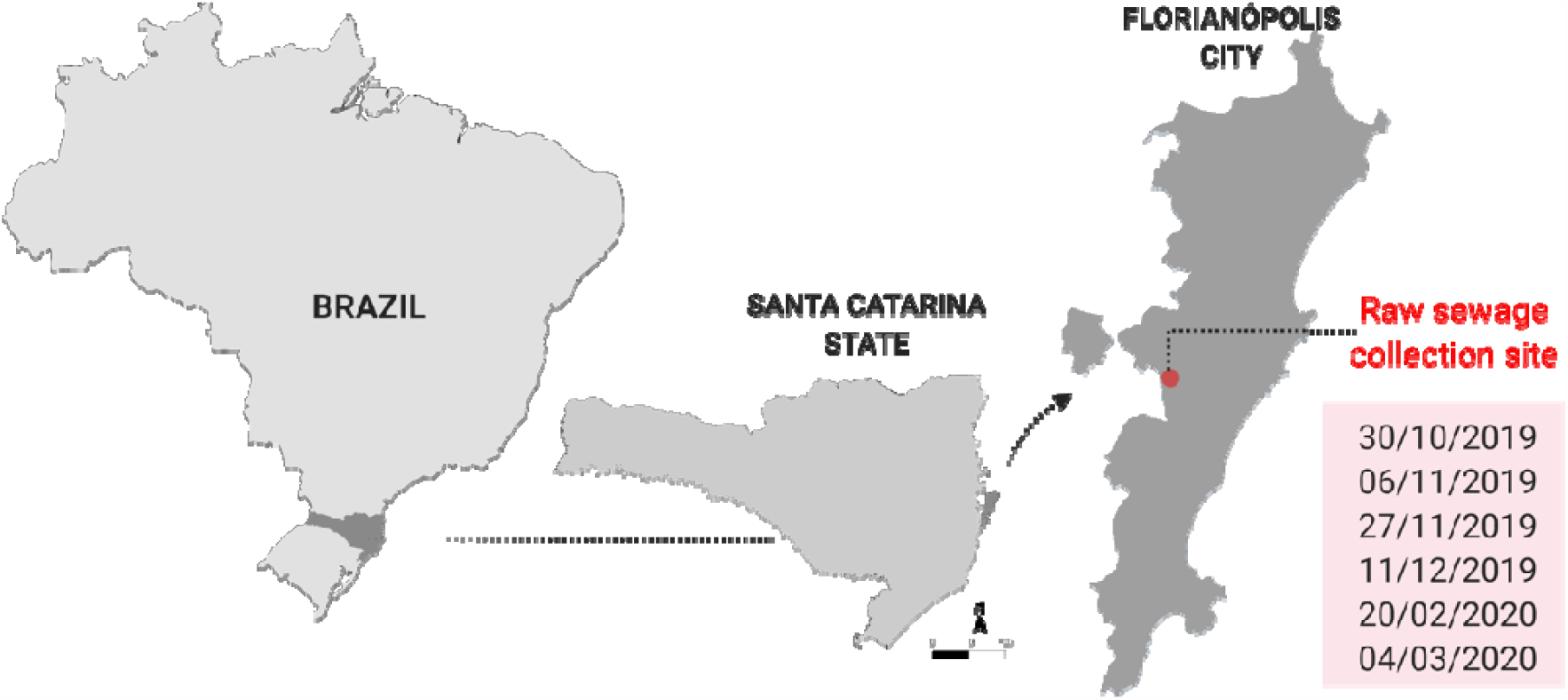
Maps of the sampling location in Florianópolis city, Santa Catarina State, Brazil.

The main physical-chemical features of urban sewages sampled are shown in Table 1. Viral RNA extraction from the sewage samples worked adequately considering the nature of the matrix, showing process efficiency values ranging from 1.6 % to 2.6 %, with an overall value of 2.1% ± 0.2% (M ± ES). Notably, while samples were negative in the first two sampling events (30^th^ October and 6^th^ November, 2019), all samples in subsequent events were positive (since 27^th^ November, 2019 until 4^th^ March, 2020) (Figure 2). The overall load was 5.83 ± 0.12 log_10_ SARS-CoV-2 genome copies L^-1^, ranging from 5.49 ± 0.02 (27^th^ November, 2019) to 6.68 ± 0.02 (4^th^ March, 2020) (Figure 2). Those SARS-CoV-2 RNA loads are similar to those found in studies performed in France [15], Spain [12], and USA [11,14]. Remarkably, SARS-CoV-2 RNA was detected as early as 27^th^ November, 2019, 66 days in advance of the first COVID-19 confirmed case in the Americas (in the USA), 91 days in advance of the first case in Brazil, and 97 days in advance of the first confirmed case in Santa Catalina Region. This demonstrates that SARS-CoV-2 was being shed within the community for several months prior to the first cases being reported by regional, national or Pan-American authorities. Few data are available on retrospective studies of SARS-CoV-2 RNA detection in sewage prior to onset of COVID-19 clinical cases in the region of the study. Randazzo et al. [12] conducted a retrospective study of wastewater process plants in the Murcia Region, Spain, and compared the observed data to declared COVID-19 cases at municipality level: the presence of SARS-CoV-2 RNA in sewage was also earlier than the first reported cases.

**Table 1.** Sewage system and physical-chemical characterization

| Samples collected | Sewer at collection point |  |  | Sewage physic-chemical characterization |  |  |  |  |
| --- | --- | --- | --- | --- | --- | --- | --- | --- |
|  | Average Flow (L.s<sup>-1</sup>) | Diameter (mm) | Population served (inhabitants) | COD <sup>a</sup> (mg L<sup>-1</sup>) | BOD <sup>b</sup> (mg L<sup>-1</sup>) | pH | Alkalinity (mgCaCO<sub>3</sub> L<sup>-1</sup>) | TSS <sup>c</sup> (mg L<sup>-1</sup>) |
|  | 1.5 | 100 mm | 5.000 | 703 | 338 | 7.9 | 320 | 142 |

<sup>a</sup> COD: Chemical Oxygen Demand
<sup>b</sup> BOD: 5 days Biochemical Oxygen Demand
<sup>c</sup> Total Suspended Solids

**Figure 2:**
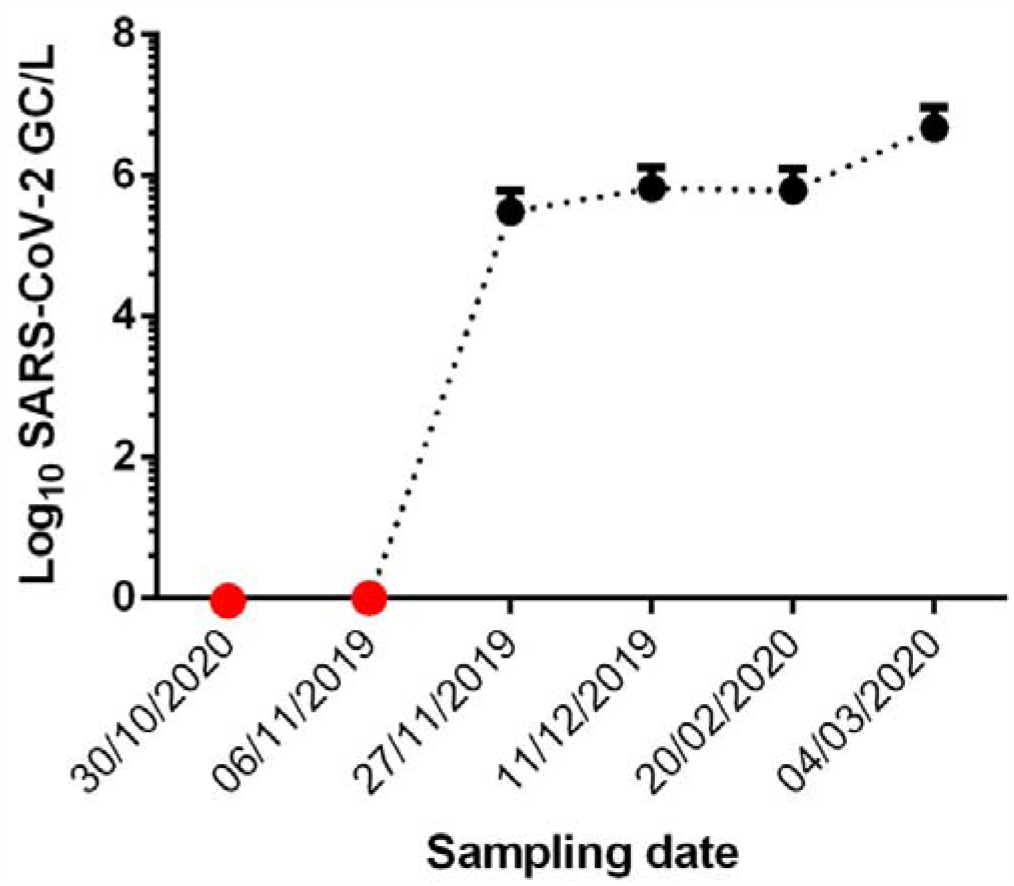
Evolution of the presence of SARS-COV-2 RNA in urban sewage, Florianopolis, Brazil. Red dots represent negative results. Black dots represent log_10_ SARS-CoV-2 genome copy per litre of urban sewage (M ± ES)

Interestingly, while the SARS-CoV-2 RNA loads were stable until late February (5.49 ± 0.02, 5.82 ± 0.01 and 5.65 ± 0.10 log_10_ SARS-CoV-2 genome copies L^-1^ in 30^th^ November, 2019, 11^th^ December, 2019 and 20^th^ February, 2020, respectively), an increase of approximately 1 log_10_ was observed on 4^th^ March 2020 (6.68 ± 0.03 log_10_ SARS-CoV-2 genome copies L^-1^), coinciding with the first COVID-19 case diagnosed in the region. Unfortunately, 10 days after the last sampling, the Federal Brazilian government ordered a national lockdown which impaired the continuation of the sampling and therefore the confirmation of that increasing tendency.

In conclusion, we have confirmed the presence of SARS-CoV-2 in the Americas as early as 27^th^ November, 60 days ahead of the reports of COVID-19 cases in the continent and more than 90 days in the case of Brazil. Therefore, our findings demonstrate that SARS-CoV-2 was circulating unnoticed in the community for some months before pandemic status was declared. Our results also show that the SARS-CoV-2 load remained constant until early March, then rose coinciding with the onset of COVID-19 cases in Santa Catalina region. Consequently, this study demonstrates that monitoring of SARS-CoV-2 in wastewater or urban sewage is an excellent tool for anticipating potential epidemiological outbreaks, and would be highly valuable in helping Public Health authorities to define protection measures.

## Data Availability

The data are available upon request

## References

1. WHO, 2020. Coronavirus disease (COVID-2019) situation reports. https://www.who.int/emergencies/diseases/novel-coronavirus-2019/situation-reports. Last access on 25th June.

2. Nsoesie EO, Rader B, Barnoon YL, Goodwin L, Brownstein JS. Analysis of hospital traffic and search engine data in Wuhan China indicates early disease activity in the Fall of 2019. Dash.Harvard.edu: 2020: http://nrs.harvard.edu/urn-3:HUL.InstRepos:42669767

3. van Doremale n N, Bushmaker T, Morris DH, Holbrook MG, Gamble A, Williamson BN, et al. Aerosol and Surface Stability of SARS-CoV-2 as Compared with SARS-CoV-1.N Engl J Med. 2020:382(16):1564–1567.

4. Ding s, Liang TJ. Is SARS-CoV-2 Also an Enteric Pathogen With Potential Fecal– Oral Transmission? A COVID-19 Virological and Clinical Review. Gastroenterol. 2020: https://doi.org/10.1053/j.gastro.2020.04.052

5. Lescure FX, Bouadma L., Nguyen D, Parisey M, Wicky PH, Behillil S, et al. 2020. Clinical and virological data of the first cases of COVID-19 in Europe: a case series. Lancet Infect. Dis. 2020: 20(6):697–706.

6. Choi PM, Tscharke BJ, Donner E, O’Brien JW, Grant SC, Kaserzon SL, Mackie R, O’Malley E, Crosbie ND, Thomas KV, Mueller JF. Wastewater-based epidemiology biomarkers: past, present and future. TraC Trends Anal. Chem. 2018: 105: 453–469

7. Prevost B, Lucas FS, Goncalves A, Richard F, Moulin L, Wurtzer S. Large scale survey of enteric viruses in river and waste water underlines the health status of the local population. Environ. Int. 2015: 79: 42–50.

8. Ahmed, W., Angel, N., Edson, J., Bibby, K., Bivins, A., O’Brien, J.W., et al. First confirmed detection of SARS-CoV-2 in untreated wastewater in Australia: a proof of concept for the wastewater surveillance of COVID-19 in the community. Sci. Total Environ. 2020: 728: 138764.

9. La Rosa G, Iaconelli M, Mancini P, Bonanno Ferraro G, Veneri C, Bonadonna L, et al. First detection OF SARS-COV-2 IN untreated wastewaters IN Italy. Sci. Total Environ. 2020: 736: 139652

10. Medema G, Heijnen L, Elsinga G, Italiaander R, Brouwer A. Presence of SARS-Coronavirus-2 RNA in Sewage and Correlation with Reported COVID-19 Prevalence in the Early Stage of the Epidemic in The Netherlands. Environ. Sci. Technol. Lett. 2020: https://doi.org/10.1021/acs.estlett.0c00357

11. Nemudryi A, Nemudraia A, Surya K, Wiegand T, Buyukyoruk M, Wilkinson R, et al. Temporal detection and phylogenetic assessment of SARS-CoV-2 in municipal wastewater. medRxiv 2020: https://doi.org/10.1101/2020.04.15.20066746

12. Randazzo W, Truchado P, Cuevas-Ferrando E, Simon P, Allende A, et al. SARS-CoV-2 RNA in wastewater anticipated COVID-19 occurrence in a low prevalence area. Water Res. 2020: 181: 115942

13. Rimoldi SG, Stefani F, Gigantiello A, Polesello S, Comandatore F, Mileto D, et al. 2020. Presence and vitality of SARS-CoV-2 virus in wastewaters and rivers. medRxiv.

14. Wu F, Xiao A, Zhang J, Gu X, Lee WL, Kauffman K, et al. SARS-CoV-2 titers in wastewater are higher than expected from clinically confirmed cases. medRxiv. 2020: https://doi.org/10.1101/2020.04.05.20051540

15. Wurtzer S, Marechal V, Mouchel JM, Moulin L. Time course quantitative detection of SARS-CoV-2 in Parisian wastewaters correlates with COVID-19 confirmed cases. medRxiv. 2020: https://doi.org/10.1101/2020.04.12.20062679

16. Viancelli A, Garcia LA, Kunz A, Steinmetz R, Esteves PA, Barardi CR. Detection of circoviruses and porcine adenoviruses in water samples collected from swine manure treatment systems. Res Vet Sci. 2012:93(1):538–54

17. Diez-Valcarce M, Cook N, Hernández M, Rodríguez-Lázaro D. Analytical application of a sample process control in detection of foodborne viroses. Food Anal. Meth. 2011: 4: 614–618.

18. Di Bartolo I, Diez-Valcarce M, Vasickova P, Kralik P, Hernandez M, Angeloni G, et al. Hepatitis E virus in pork production chain in Czech Republic, Italy, and Spain, 2010. Emerg. Inf. Dis. 2020: 18: 1282–1289.

19. CDC. CDC 2019-novel coronavirus (2019-nCoV) real-time RT-PCR diagnostic panel. https://www.fda.gov/media/134922/download.

20. Chan JFW, Yuan S, Kok KH, To KKG, Chu H, Yang J, et al. Jasper Fuk-Woo Chan - A familial cluster of pneumonia associated with the 2019 novel coronavirus indicating person-to-person transmission: a study of a family cluster. Lancet.. 2020: 395(10223):514–523.

